# A systematic review of previous MRI studies demonstrating the relationship between season and brain volume

**DOI:** 10.1101/2022.02.17.22271139

**Authors:** Naif A. Majrashi, Ali S. Alyami

## Abstract

**Introduction:** Specific diseases such as Alzheimer’s, schizophrenia and multiple sclerosis have been associated with environmental changes, including changes in season. The relationship between changes in season or photoperiod and brain volume has been widely studied in animals. However, the relationship between changes in season and brain volume in humans is not yet well established. Here, we aim to provide a comprehensive and systematic review of magnetic resonance imaging (MRI) studies examining the effects of changes in season or photoperiod on brain volume.

**Methods:** We used a systematic review approach to study reports on the relationship between changes in season or photoperiod and brain volume using MRI. PubMed database and Google Scholar search engines were used to locate appropriate. PRISMA was used in selection of appropriate studies on season or photoperiod and brain volume using MRI.

**Results:** Five studies were included in the current review: three examined the relationship between changes in photoperiod and brain volumes while two studies examined the relationship between changes in season and brain volume.

**Conclusion:** When studying other variables like changes in temperature and humidity on how they affect brain volume, no effect was observed. It was, however, observed from studies that hydration status changed brain volumes when measured using MRI. Overall, we found evidence demonstrating differences in human brain volumes across different seasons. We suggest future longitudinal research to prove changes in brain volume across different seasons.

## Introduction

MRI is deemed to be a significant diagnostic tool for many diseases in the body (Berger, 2002). It has become an important tool for brain volume estimation, which is considered significant for measuring normal and abnormal changes in the brain, due to excellent levels of image spatial resolution and between-tissue contrast through the reliance on various MR image contrasts. Previous MRI studies have provided key insights into macro- and micro-structures of the brain (Walhovd, Johansen-Berg & Káradóttir, 2014). For example, T1 weighted MRI studies, which have quantified brain volumes analysed on voxel-wise and volume-segmentation bases across the whole brain and substructures, have highlighted brain volume changes associated with several factors such as aging, and that with increasing age the brain decreases in overall volume (Raz et al., 1997, 2015). Previous diffusion tensor imaging (DTI), a technique used to estimate the axonal or white matter organisation in the brain, MRI studies have also played a prominent role in the characterisation of white matter changes in the brain (Bennett & Madden, 2014).

There are two prominent adaptive hypotheses, the Expensive Brain Hypothesis (EBH) and Cognitive Buffer Hypothesis (CBH) proposed to explain brain evolution. CBH proposes that brain size increases with seasonality, and this increases cognitive ability of species. EBH on the other hand assumes brain size decreases with seasonality (Jun Gu et al., 2017). A previous study by Pyter et. al, (2007) has found that short days reduced brain size, impaired hippocampal-based spatial learning and memory, reduced the hippocampal volume, and decreased the CA1 hippocampal spine density in adult male white-footed mice. Studies on effects of season on human body have found a relationship between changes in environmental factors and diseases. Specific diseases like Alzheimer’s, schizophrenia and multiple sclerosis have been associated with environmental changes, including change in altitude. Although the brain’s hippocampal volume changes associated with learning and exercise have been studied, natural environmental effects have not been intensively explored.

Previous studies investigating the relationship between seasonal or photoperiodic changes and brain volume have been conducted in in birds and non-human mammals with little studies in humans (Book et al., 2021; Miller et al., 2015). Changes in season have been observed especially in the hippocampus and amygdala, reflecting seasonal neuroplasticity (Clayton, Reboreda & Kacelnik, 1997; Pyter, Reader & Nelson, 2005; Sherry & Hoshooley, 2010; Walton et al., 2011; Workman et al., 2011; Yaskin, 2011). White-footed mice Peromyscus leucopus, exhibit smaller hippocampal volume during the winter season compared to the autumn or summer seasons. According to Book et al., (2021), the relationship between weather or seasonal changes and brain volume can best be studied using a large data set. This study used healthy subjects in a 15-year period, studying the effects of different seasons on brain volume at Hartford CT USA. Other variables studied in this study included the change in the hippocampal volume in relation to seasonal changes. A previous study (Majrashi, Ahearn & Waiter, 2020) has also studied the brainstem volume against mood and depression in seasonal variations in large dataset. This study found that there was a positive relation between photoperiod and brainstem volumes. Another cross-sectional studies by (Majrashi et al., 2020; Miller et al., 2015) have also found a positive correlation between changes in photoperiod and hippocampal volume in both females and males. Although seasonal changes in brain volumes have been widely studied in animals, seasonal differences nor changes in human brain volumes have not yet been established well. Here, we aim to provide a systematic review of cross-sectional and longitudinal MRI studies that have examined the effects of changes in season or photoperiod on human brain volume. Investigation of brain volume differences across seasons with different photoperiod may improve understanding of naturally occurring neuroplasticity. It might also increase our understanding of the temporal factors, such as duration of day length, that play a significant role in shaping naturally occurring neuroplasticity. In light of research demonstrating the influence of seasonal differences in photoperiod on brain volumes in human, we hypothesise that brain volumes such as the hippocampus and amygdala are increased with during the summer months with longer photoperiod days and decreased during the winter months with shorter photoperiod days.

## Methods

### Justification

Several previous studies have been conducted on effects of daylight on different parts of the brain while few studies have taken into consideration how weather conditions affect the brain volume (Book et, al, 2021). The current study used a systematic review approach to study reports on the relationship between changes in season or photoperiod and brain volume using MRI, with the aim to explore the relationship between seasonal changes and brain volume.

## Objectives

To explore the relationship between changes in season or photoperiod and brain volume.

### Data source

Online Med Pub database and Google Scholar search engines were used to locate relevant literature for this study. Other relevant studies were manually searched with guidance from refences under selected studies whose abstracts were further read for use in the study.

### Study selection

The authors independently used the PRISMA flow diagram below (Figure 1) to select appropriate studies on season or photoperiod and brain volume using MRI. For studies to be included in the review, the following criteria had to be met:

1. Initially the search using search terms “Season or photoperiod and brain volume” yielded 283 results. Another term MRI was included in the search and 34 results were yielded. In the MeSH the word full text was selected and a search done which yielded 33 results.
2. Abstracts of these 33 reports were downloaded and studied to further exclude the studies that were not addressing the topic and 5 studies were found to be relevant.
3. Studies were screened by reading their titles and abstracts relevant to season or photoperiod and brain volume using MRI were selected.
4. Full texts of selected studies were downloaded for in-depth reading and extraction of relevant information as per the topic.
5. Inclusion criteria: all relevant published and unpublished reports on season and brain volume, volume of brain regions or brain parts studied using MRI and in english language were selected.
6. Exclusion criteria: studies on brain volume, brain parts and other seasons like sports seasons and injuries were excluded.

**Figure 1.**
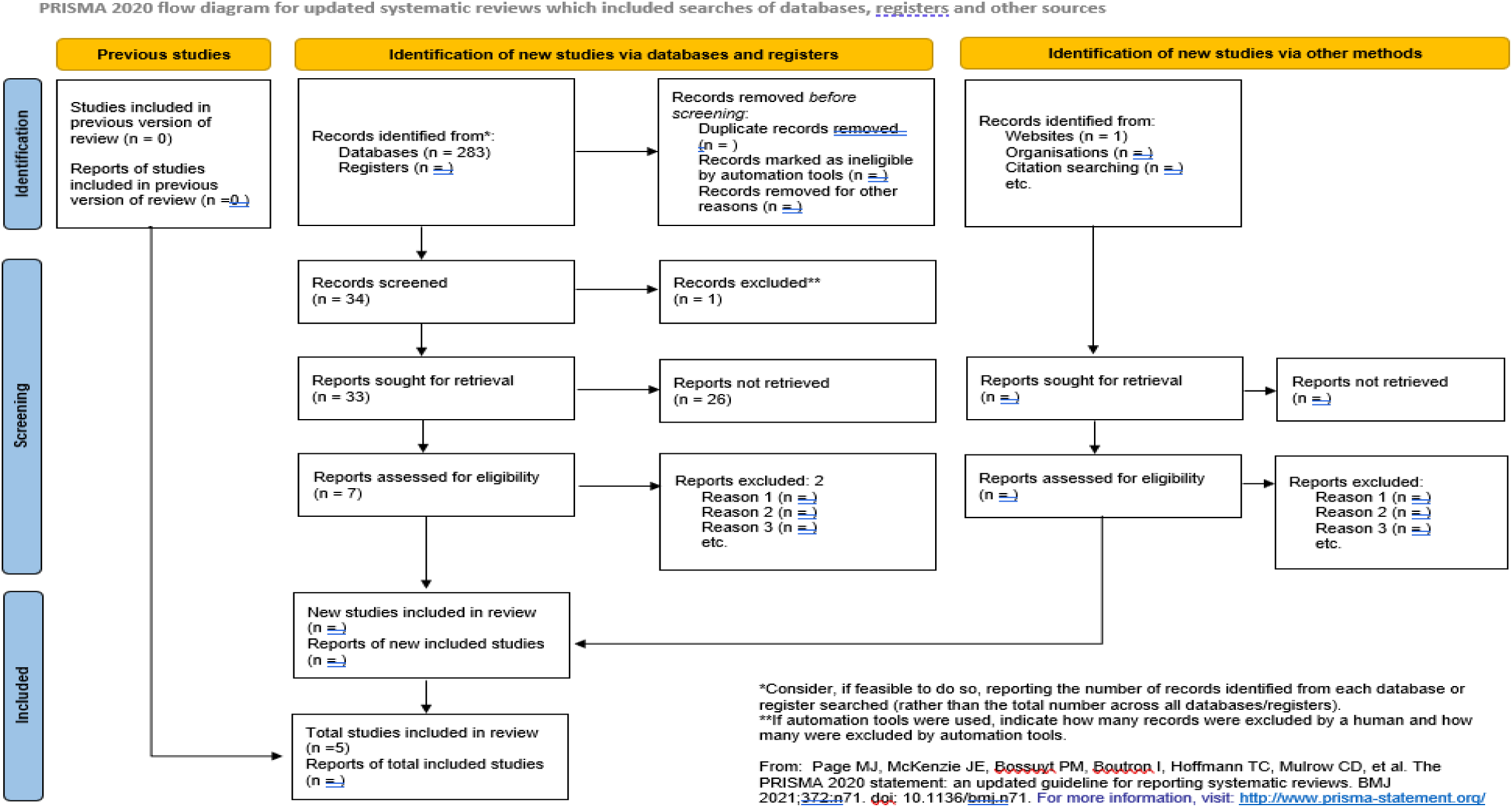
PRISMA flow diagram used for selection of reports

## Data extraction and analysis

### Volumetric Segmentation and Analyses

Most MRI studies used the volumetric segmentation, and it was completed using available software packages at different centres in studying brain volume or different parts of the brain against specified parameters. The two authors independently extracted the following details using a structured data abstraction form: aspect of brain region examined and the processing tool.

### Voxel-based morphometry (VBM) Analysis

Voxel-based morphometry analysis was performed in some studies discussed in the report and analysis used available software and images were screened for artifacts and anatomical abnormalities. MR images were then segmented in white matter, grey matter, and cerebrospinal fluid by using the available segmentation toolbox in available software.

## Results

### Study selection

The PRISMA chart (figure1) was deployed in screening two hundred and eighty-three (283) titles and their abstracts for analysis. Thirty-three (33) studies were further selected for review with extensive reading of their abstracts. Five (5) studies were further selected as appropriate for the current study (Table 1).

**Table 1.** MRI data from previous studies investigating the relationship between season or photoperiod and brain volume

| Author | No. of participants | MRI Scanner | MRI Design | Parameter studied | Significant results |
| --- | --- | --- | --- | --- | --- |
| Book et al. (2021) | 1,779 female and 1,500 male | 3T Siemens Skyra | T1WI | Weather and season vs human brain volume | Significant |
| Majrashi et, al (2020) | 9289 (51.8% females, 48.1% males) | 3T Siemens Skyra with a standard Siemens 32-channel RF receive head coil43. | T1W-MPRAGE | Photoperiod vs brainstem volumes | Significant |
| Majrashi et, al (2020) | 10,033 participants aged 37–73 years (from 2006 – 2010) | 3T Siemens Skyra | T1W-MPRAGE | Sex differences in the association of photoperiod with hippocampal subfield volumes | Significant |
| Pantazatos (2014) | 550 normal, healthy adults | Philips 1.5T, General Electric 1.5T, Philips 3T | T1WI | GM, white matter [WM], and cerebrospinal fluid) and Season of birth (SOB) | Significant |
| Miller et al. (2015) | 404 healthy subjects | 3T Trio TIM whole-body scanner (Siemens, Erlangen, Germany) | T1W-MPRAGE | Hippocampal volume vs photoperiod | Significant |

T1-weighted magnetization prepared rapid gradient echo imaging (MPRAGE)

All studies employed a cross-sectional MRI design and primary analyses were significant for all studies. Three studies (Miller et al., 2015; Majrashi, Ahearn & Waiter, 2020, 2020) found that participants exposed to longer photoperiod days exhibited larger hippocampal and brainstem substructure volumes compared to those exposed to shorter photoperiod days. Further, (Pantazatos, 2014) found significant annual periodicity in the left superior temporal gyrus (STG) and grey matter volume, with a peak towards the end of December. Finally, (Book et al., 2020) found positive relationships between left and right cerebellum cortex and white matter between seasons (the comparisons of January/June and January/September) and negative relationships between several subcortical ROIs for the summer months compared to January.

## Discussion

The current systematic review is the first to review brain volume associations with changes in season or photoperiod in humans. The systematic review has reviewed and summarized all previous studies investigating the relationship between changes in season or photoperiod and brain volumes. In brief, previous studies (Majrashi et al., 2020; Miller et al., 2015) found a significant correlation between photoperiod and total and hippocampal subfield volumes corrected for age and total brain volume within a large population cohort. Hippocampal and subfield volumes were found to be larger during summer season (longer photoperiod days) and smaller during winter season (shorter photoperiod days). It also found sex differences, which were confined to the left side, in the association of photoperiod with hippocampal subfield volumes, with females showing greater rate of change compared with males. Further, it found that dentate gyrus (DG) and Connu Ammonis (CA1) hippocampal subfields in both hemispheres have the highest rate of change. These studies showed that 0.7% of the variation in hippocampal volume was accounted for by variations in photoperiod on the day of scan. In addition, a previous study (Majrashi, Ahearn & Waiter, 2020) demonstrated that photoperiod was negatively associated with low mood and anhedonia depressive scores in females but not in males, where longer photoperiod days were associated with reporting reduced low mood and anhedonia. Further, this study has found that brainstem substructure volumes fully mediate the seasonality of depressive symptoms in females but not in males, suggesting that photoperiod may be an important environmental factor to consider when studying brain volumes. This study lays a base in exploring further the relation between season and brain volume, as this concept has not been explored much. Moreover, a previous study by Pantazatos (2014) explored season of birth effects (SOB) on human brain volume. The study found out that some environmental variables (temperature and humidity) were associated with changes in the human brain structure, and this persisted through adulthood. This study also found out that the overall effect of SOB was more on males than females in the left STG, exhibiting reduced grey matter volume in summer or spring relative to winter or fall births. Variables like physical activity and sleep duration were associated with hippocampal volume changes. The findings from these previous studies advance our understanding of the hippocampal and brainstem morphologies and suggest that hippocampal and brainstem may be an important neural substrate in the pathophysiology of seasonal mood disorders. These finding point to the evidence supporting the role of photoperiod on brain structural plasticity.

We have found two brain regions including the hippocampus and brainstem to be associated with changes in season and photoperiod. The brainstem controls the vital body functions such as sleep or wake processes, blood pressure (Brennan et al., 1982), heart rate, and cardiac and respiratory (Kristal-Boneh et al., 2000), while the hippocampus is responsible for forming and retrieval memory and controlling emotional processes, respectively. It has been proposed that decreased brain volumes including the hippocampus during the winter may be a result of decreased need for spatial memory in foraging behaviours (Pyter, Reader & Nelson, 2005). Therefore, this evidence supports the notion that the brain functions are also affected by seasons. This may suggest that hippocampal and brainstem functions may be more vulnerable to seasonal differences, and that their functions may be impaired in winter and improved in summer.

### Avenues for future research

One interesting avenue for research would be conducting a longitudinal study, with healthy participants, measuring brain volumes in both summer and winter seasons to determine within-individual seasonal hippocampal, amygdala and brainstem volumes change. Another interesting avenue for research would be replicating the results of seasonal variations in brain volumes, particularly the hippocampus, in a sample with higher depression scores, suggesting that hippocampal volumes would also mediate the seasonality of depressive symptoms. Looking at other brain areas important to mood changes which may also exhibit seasonal plasticity would be important for future research. Previous studies found seasonal changes in hypothalamus, thalamus, neocortex and striatum in common shrew (Lázaro et al., 2018; Puga et al., 2018), brain regions which have also been implicated in mood and the development of depression (Price & Drevets, 2010; Heshmati & Russo, 2015). Therefore, photoperiod may affect the morphology of such brain regions and may be important to include photoperiod in the relationship between depressive symptoms and volumes of such brain regions. One more interesting avenue for research would be that associations between cognitive measures and brain volumes including the hippocampus and brainstem may suggest the importance for controlling for photoperiod in future studies. It has been shown that IQ scores (cognitive subjective decline scores) are negatively associated with hippocampal and brainstem volumes (Amat et al., 2008; Yue et al., 2018; Valdés Hernández et al., 2016; Ikram et al., 2010). Therefore, it may be important to include photoperiod in the longitudinal relationships between hippocampal and brainstem volumes and IQ or cognitive scores.

## Conclusion

Studies discussed in this paper suggest that photoperiod could be an important variable to analyse in relation to brain volume. Data presented in the study offer preliminary indication that human hippocampal plasticity may be associated with photoperiod hence a need for longitudinal studies. Many studies have been conducted on effects of daylight on different parts of the brain while few studies have taken into consideration how weather conditions affect the brain volume, hence the need to explore in depth the effects of seasonal changes in the brain volume.

## Data Availability

All relevant data are within the manuscript and its Supporting Information files

NA

## Acknowledgments

This research is supported by Jazan University, Saudi Arabia.

## Registration and protocol

The current systematic review was not registered.

